# Thymus Composition, Disease Control, and Toxicity in Locally Advanced Lung Cancer

**DOI:** 10.1101/2025.10.20.25338395

**Authors:** Tafadzwa L. Chaunzwa, Abraham J. Book, Biniam Garomsa, Yuzhong J. Meng, Elaine Yang, Gokul Krishnan, Alexis Chidi, Ryan Kim, Jennifer Ma, Gregory J. Riely, James Huang, Charles B. Simone, Narek Shaverdian, Daniel R. Gomez

**Author notes:** Co-first authors (Tafadzwa L. Chaunzwa, Abraham J. Book). Corresponding author: Tafadzwa L. Chaunzwa, Assistant Member, Department of Radiation Oncology, Memorial Sloan Kettering Cancer Center; Advanced Computing and Oncology Laboratory, Mortimer B. Zuckerman Research Center, 417 East 68th Street, New York, NY 10065.

## Abstract

Thymic involution, characterized by adipose replacement of functional thymic tissue, is a broadly recognized feature of age-related immunosenescence. Currently, no established, non-invasive method measures residual thymus in adults, its relationship with host immunity, or its influence on tumor immunosurveillance, cancer outcomes, and treatment-related toxicities. In a multi-institutional cohort of patients with locally advanced non-small cell lung cancer (NSCLC), we created a new CT-based metric to estimate residual functional thymic tissue and examined its association with toxicities and clinical outcomes. We defined a thymic region atlas and employed a novel segmentation model that integrates Transformer-based attention with convolutional detail preservation to automatically detect and delineate the thymic region on chest CT images. We then computed a new radiographic parameter, capturing the proportion of persistent thymic tissue or “percent thymic tissue” (pTT), using Gaussian mixture modeling of the components of the delineated primal thymic space. We applied our framework to a publicly available NSCLC radiotherapy dataset (MAASTRO) and an institutional cohort at Memorial Sloan Kettering Cancer Center (MSKCC), assessing associations with age and sex. Kaplan-Meier estimates and Cox proportional hazards modeling were used to evaluate the association between pTT and distant metastasis-free survival (DMFS), locoregional failure (LRF), progression-free survival (PFS), and overall survival (OS). A total of 464 patients with stage III NSCLC, pre-treatment chest CT scans, curated clinical data, and long-term follow-up were analyzed. Of these, 277 were from MAASTRO and 187 from MSKCC. Our pTT method allowed tissue decomposition without manual thresholding or shape priors and was inherently robust to segmentation variability. pTT decreased with age at a rate of 0.36 percentage points per year (p < 0.0001) in the MAASTRO cohort and 0.40 percentage points per year in the MSKCC group. pTT was significantly lower in males (MAASTRO: 9.75% vs. 17.0%, p<0.001; MSKCC: 5.97% vs. 12.3%, p<0.001). On multivariable Cox regression adjusting for age, sex, and histology, pTT was an independent predictor of survival (MAASTRO: aHR = 0.71, 95% CI: 0.50-0.99; p = 0.049; MSKCC: aHR = 0.51, 95% CI: 0.27-0.95; p = 0.058). Notably, pTT was also associated with DMFS (aHR = 0.51, 95% CI: 0.27-0.95; p = 0.03), LRF (aHR, 0.96; 95% CI, 0.93–0.99; p = 0.014), and PFS (aHR = 0.57, 95% CI: 0.32-1.01; p = 0.052) in the MSKCC cohort. There was a link between pTT and 12- and 24-month estimates for all end-points. pTT remained a significant predictor of improved DMFS even after adjusting for thymic radiation dose (aHR = 0.97; 95% CI, 0.94–0.99; p = 0.037), which itself was independently associated with inferior DMFS (aHR = 2.01; 95% CI, 1.02–3.99; p = 0.045). The LRF association also persisted after adjusting for thymus V20 (aHR, 0.96; 95% CI, 0.92–0.99; p = 0.008). Severe pneumonitis (grade ≥3) occurred more frequently in patients with low pTT values (13.9% vs. 5.2%; p = 0.04). Patients with both high lung V20 and low pTT represented the subgroup at greatest pneumonitis risk (20.8%, n = 11), whereas those with low lung V20 and high pTT had few observed events (1.9%, n =1). pTT also predicted durvalumab discontinuation due to treatment-related adverse events, particularly among patients with unresectable stage IIIC NSCLC (AUC = 0.8). Thus, the parameter pTT offers a reliable and interpretable non-invasive quantitative measure of residual functional thymus in adults, reflects age-related thymic involution, and independently predicts survival and treatment-related toxicity in stage III NSCLC. These findings support pTT as a structural imaging indicator of thymic function and suggest its potential for studying how cancer therapies impact host immunity, which may, in turn, influence long-term lung cancer treatment outcomes.

## Introduction

Thymic involution – marked by the gradual replacement of functional thymic epithelial tissue and mesenchymal stroma with fibrous and adipose tissue – is a well-described and widely recognized hallmark of immunosenescence that reflects the gradual decline of structural immune function with age^1–4^. Although long viewed as evidence that the thymus is vestigial in adult physiology, residual thymic tissue and function persist well into adulthood^5,6^ and may be especially critical during periods of physiological stress or following therapeutic injury^4,7^. Furthermore, patient registry data recently reported in the New England Journal of Medicine associated adult thymectomy with increased risk of various human illnesses, including cancer, as well as higher mortality, sparking broad interest in reevaluating the role of the thymus in adult patients^8^.

In cancer, mainstay treatments such as chemotherapy and radiation therapy can accelerate thymic remodeling through direct damage to the tissue or systemic effects^7^. Yet, the relationship between therapeutic exposure, thymic structural decline, and clinical outcomes is incompletely understood in clinical oncology^9,10^. Moreover, in the modern era, where immunotherapies targeting key immune checkpoint pathways (e.g., PD-1/PD-L1, CTLA-4, LAG-3, TIGIT) have become integral to the standard treatment for locally advanced and metastatic non-small cell lung cancer (NSCLC)^11–16^, maintaining host capacity to generate a strong antitumor immune response has become increasingly important. Damage of immune organs such as the thymus may blunt the effects of immune checkpoint inhibitors (ICIs)^17,18^. Additionally, the clinical observation of infrequent but profound responses to ICIs in patients treated for NSCLC has generated interest in developing predictive indicators in this setting^12,13,19^.

The assessment of thymic composition *in vivo* offers a unique avenue to study this often-overlooked primary lymphoid organ in the context of cancer therapy. Analyzing clinical imaging is a particularly effective exploratory method because non-invasive imaging like X-ray computed tomography (CT) of the chest is routinely performed for patients diagnosed with and treated for NSCLC across tertiary and community care settings^14,20^. However, existing methods for evaluating thymic integrity on CT have relied predominantly on morphometric surrogates or simplistic attenuation thresholds that fail to capture the heterogeneous tissue composition of the gland ^21–24^. On the other hand, while deep learning-based approaches have advanced medical image analysis, they often generate abstract features with limited interpretability and uncertain biologic significance, limiting their clinical and translational utility^25–28^. When it comes to health-related decision, patients and clinicians consistently express a preference for transparent and readily explainable decision-support systems they can understand and trust^29–31^.

In this study, we developed an interpretable, biologically grounded, CT-based model to accurately measure thymic atrophy, map the thymic microenvironment, and derive a non-invasive indicator of its structural integrity. By performing voxel-wise analysis of CT attenuation values in the primal thymic space using Gaussian mixture modeling ^32,33^, we derived a biologically meaningful, volume-independent parameter that quantifies the proportion of residual functional thymic tissue relative to adipose content. This metric, the “percent thymic tissue” (pTT), builds upon published and open-source concepts and methods^3,8,18,22,24,32,34–39^, notably a quantitative framework introduced by Okamura et al^38^, with significant mathematical refinement, to allow for standardized assessment of thymic composition across populations. We hypothesized that this radiographic marker for thymic tissue composition reflects host immunity and that, despite structural decline in adulthood, the atrophied thymus remains relevant in patients receiving treatment for locally advanced NSCLC.

We applied our pTT framework to a transatlantic, multi-institutional cohort of patients with locally advanced NSCLC treated with curative-intent concurrent chemoradiotherapy, some of whom also received adjuvant ICIs^11,14,16^. We identified pTT associations with end points including distant metastasis-free survival (DMFS), locoregional failure (LRF), progression-free survival (PFS), and overall survival (OS). We also found an association between pTT and severe pneumonitis or radiation pneumonitis, and among patients with more advanced locoregional disease, pTT predicted clinically significant, treatment-emergent adverse events (AEs) leading to discontinuation of consolidation durvalumab. Our findings demonstrate the feasibility of reliably tracking host immunity through non-invasive, biologically explainable, quantitative image-based analysis of immune organs. They also highlight the clinical significance of the thymus and its potential impact on lung cancer outcomes across various treatment modalities, including chemotherapy, radiotherapy, and immunotherapy.

## Results

### Clinical cohorts and patient characteristics

We analyzed CT images and clinical outcomes data for 464 patients with stage III NSCLC from two independent cohorts treated at cancer centers in the Netherlands and United States: the Maastricht Radiation Oncology^40^(MAASTRO) cohort (n = 277) and an Memorial Sloan Kettering Cancer Center (MSKCC) institutional cohort (n = 187). Median age in the MAASTRO cohort was 66 years (range, 34–88) and in the MSKCC cohort was 69 years (range, 45–87). Median OS was 17.9 months in MAASTRO and 35.8 months in MSKCC. Of the combined dataset, roughly 60% were male. The pTT metric was extracted from the baseline CT scan for each patient (Figure 1A). Comparable positively-skewed pTT value distributions were observed in both cohorts, with median values of 11.6% and 8.14% for MAASTRO and MSKCC, respectively (Figure 2A).

**Figure 1.**
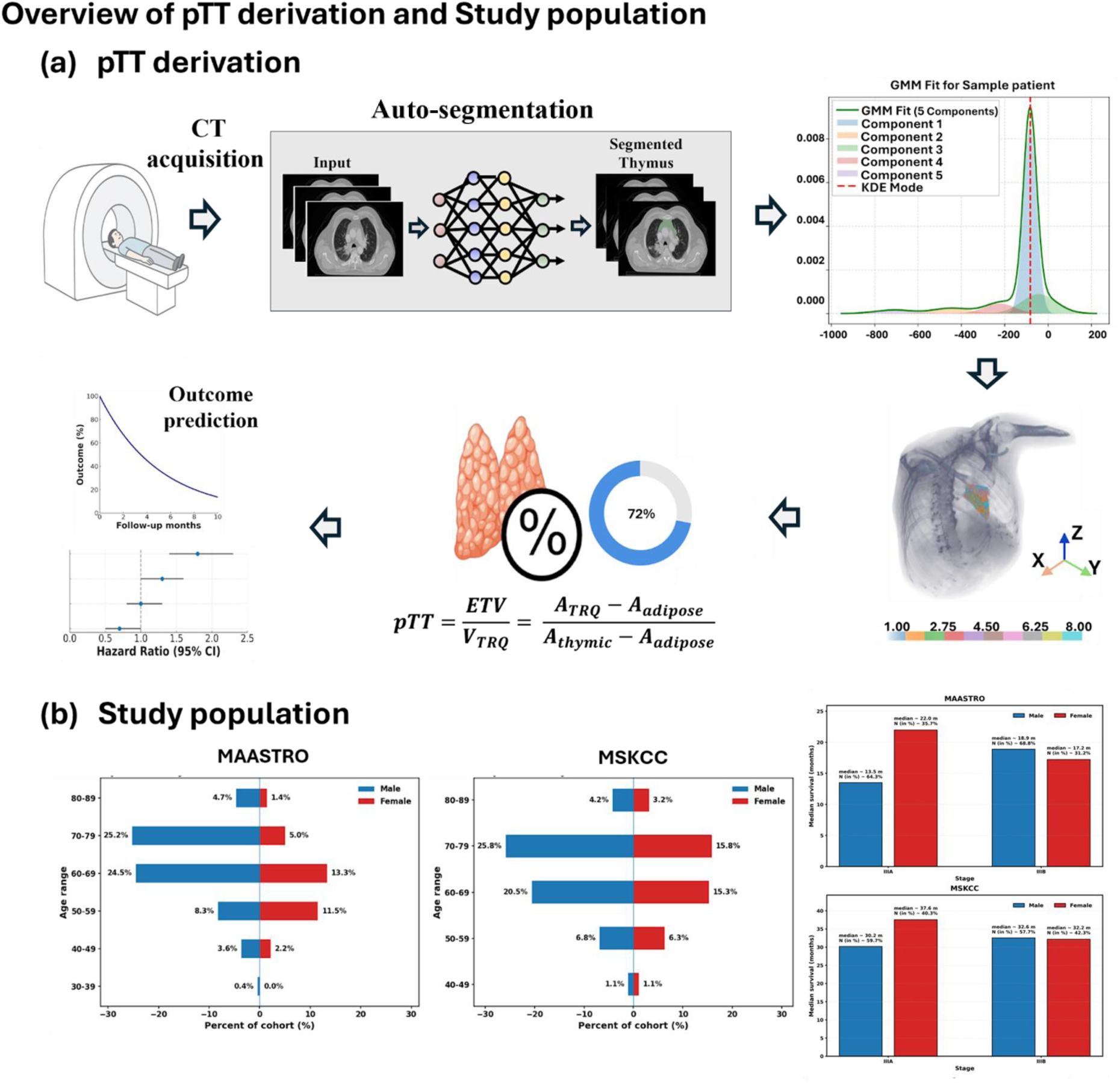
Overview of pTT extraction and application as a prognostic indicator in lung cancer. (a) pTT derivation pipeline, (b) overview of the characteristics of study population. CT: computed tomography; SWIMMER: Swin-based Immune & Malignant structure Mapping with Entropy-based Regression; GMM: Gaussian mixture modeling; pTT: percent thymic tissue; ETV: estimated thymic volume; Vtrq: volume of thymic region for quantification

**Figure 2.**
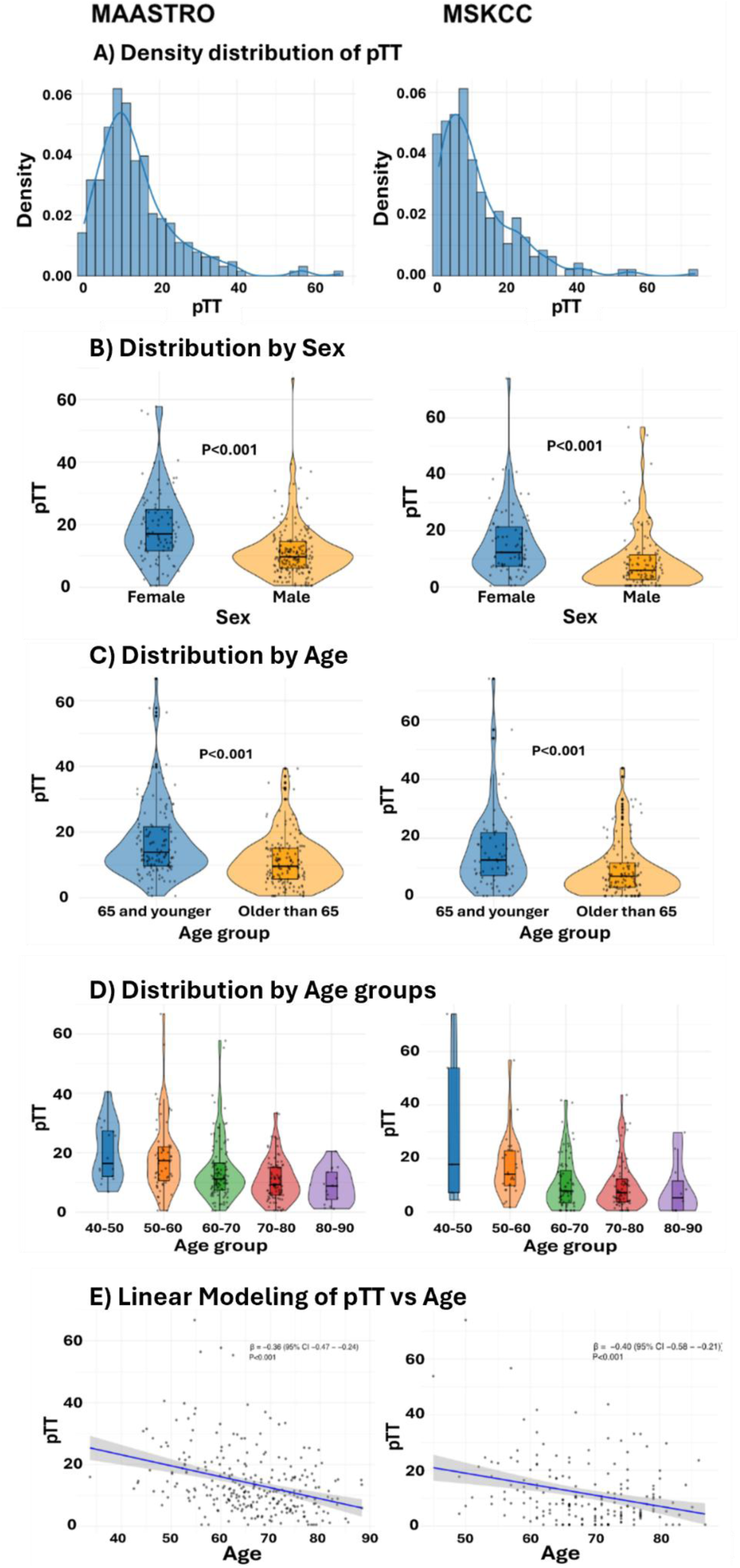
Density distribution of pTT in MAASTRO and MSKCC cohorts, and association with Age and Sex.

### Association of pTT with age and sex

Significant differences in pTT were observed across sex and age in both cohorts. Median pTT was significantly lower in males than in females in both the MAASTRO cohort: 9.75% vs 17.00%, p<0.001 and the MSKCC cohort: 5.97% vs 12.30%, p<0.001 (Figure 2B). Patients aged 65 years or younger had higher median pTT values than those older than 65 years in both the MAASTRO (13.85% vs 9.56%, p < 0.001) and MSKCC (12.60% vs 7.15%, p < 0.001) cohorts (Figure 2C). A one-year increase in age was associated with a 0.36 percentage-point decrease in pTT (β = –0.36; 95 % CI: –0.47 to –0.24; p < 0.001) in the MAASTRO cohort and a 0.40 percentage-point decrease (β = –0.40; 95 % CI: –0.58 to –0.21; p < 0.001) in the MSKCC cohort (Figures 2D and 2E). Overall, similar associations of pTT with age and sex were observed in each cohort.

### Prognostic value of pTT

We assessed the prognostic value of pTT using a subset of MAASTRO as the discovery cohort. All of these patients received chemoradiation alone for stage III NSCLC. Among the 277 patients analyzed, those with pTT ≥ 5.59% demonstrated significantly better survival outcomes compared to those with lower pTT on Kaplan-Meier analysis, with median OS of 20 months for the high pTT group compared to 17 months in the low pTT group (log-rank P = 0.02) (Figure 3A). The survival curves diverged early and remained separated throughout the follow up period. On multivariable Cox-proportional hazards analysis, adjusting for all available covariates including age, sex, and histologic subtype, pTT remained an independent predictor of OS (adjusted hazard ratio [aHR] = 0.71, 95% CI: 0.50-0.99, p=0.049) (Figure 3B). Notably, pTT remained significantly associated with survival outcome in the adjusted model whereas age did not reach statistical significance (Figure 3B).

**Figure 3.**
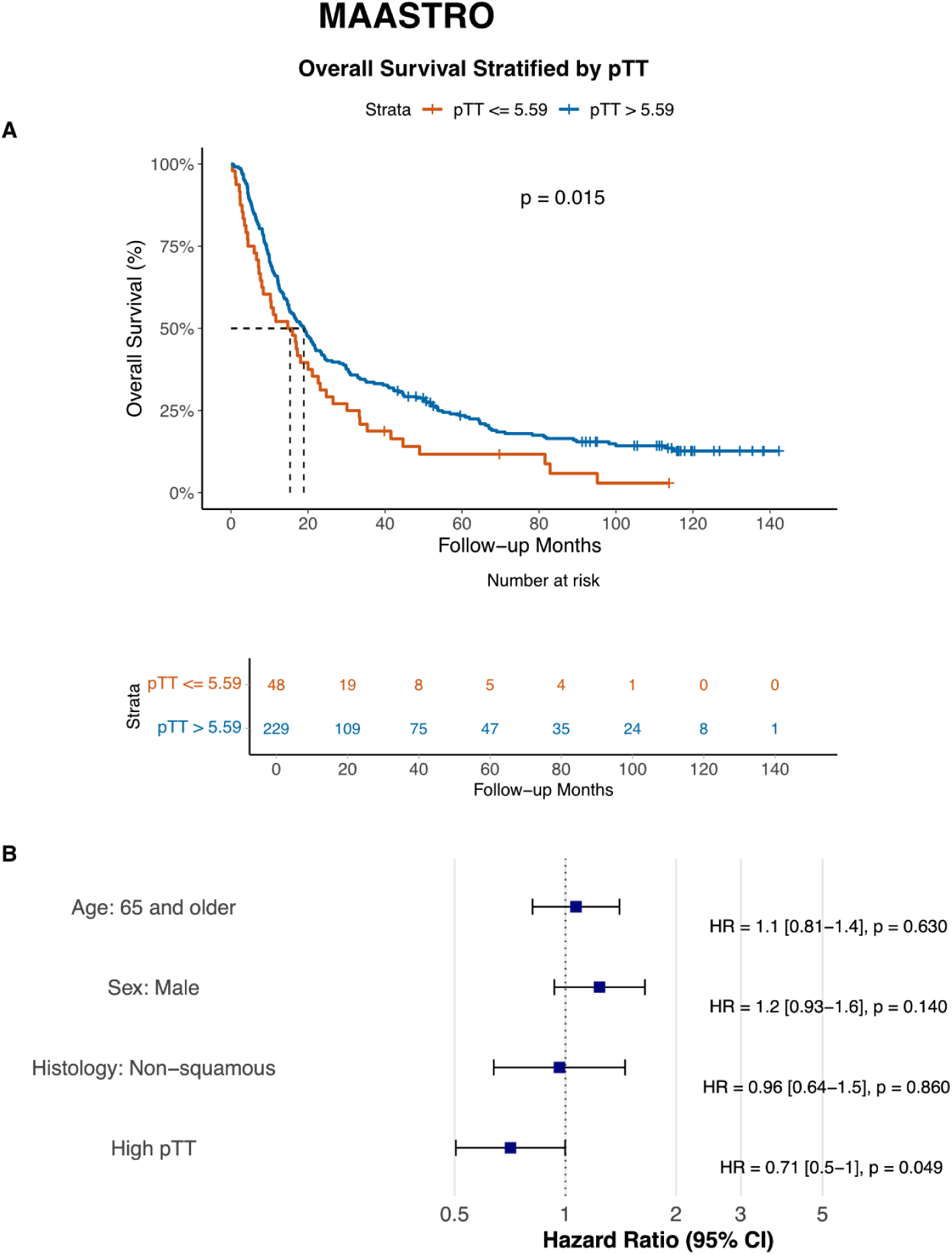
Kaplan-Meier estimates and Cox Proportional Hazard Models for the Association of pTT with overall survival (OS) in the MAASTRO cohort.

### Validation of pTT prognostic utility

A cohort of 187 consecutive patients with unresectable locally advanced NSCLC treated with chemoradiotherapy and consolidation durvalumab at MSKCC was used for independent validation and cross-modality assessment of the generalizability of pTT as a prognostic biomarker. In addition to baseline chest CT scans, all patients in the MSKCC cohort had extended clinical follow-up, tracking additional clinical end-points including DMFS and PFS. Median DMFS was not reached, median PFS was 35.3 months.

Our Kaplan-Meier analysis demonstrated that higher pTT values were associated with improved outcomes across all studied endpoints. Patients with higher pTT (above a 14-17% threshold) demonstrated improved DMFS, PFS, and OS compared to those with lower pTT (Figure 4). The specific optimal pTT threshold for stratification varied slightly for the different endpoints (pTT ≥ 14.97% for DMFS, pTT ≥ 17.23% for PFS, and pTT ≥ 14.49% for OS). Multivariable Cox models adjusting for age, sex, histology, and smoking history confirmed pTT as an independent predictor of DMFS (aHR = 0.51, 95% CI: 0.27-0.95, p=0.03), PFS (aHR = 0.57, 95% CI: 0.32-1.01, p=0.053) and OS (aHR = 0.51, 95% CI: 0.27-0.95, p=0.058). pTT remained an independent predictor in adjusted Cox models while age did not reach statistical significance for DMFS and PFS (Figure 4A and 4B).

**Figure 4.**
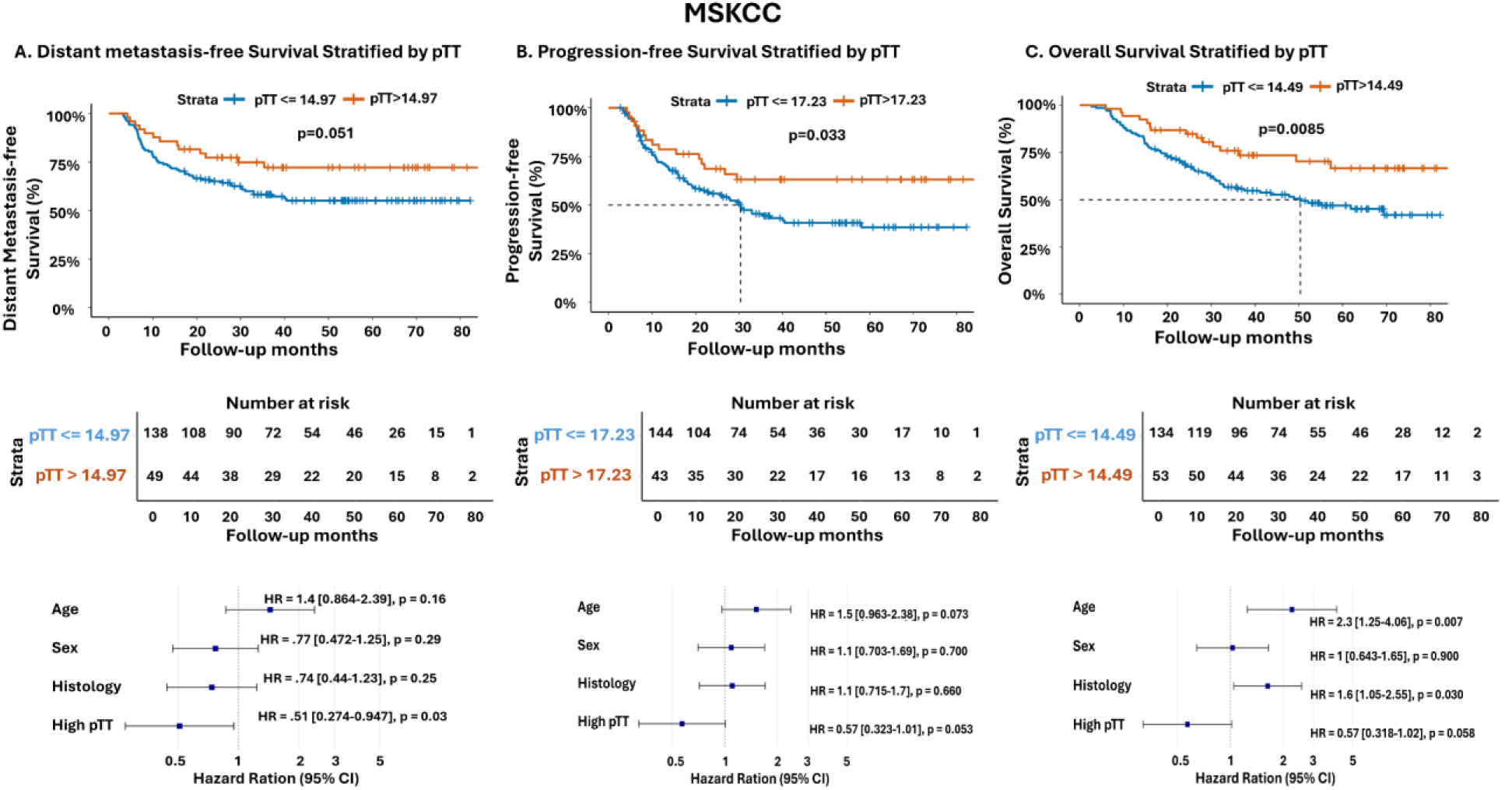
Kaplan-Meier estimates and Cox Proportional Hazard Models for the Association of pTT with distant metastasis-free survival (DMFS), progression-free survival (PFS), and overall survival (OS) in the MSKCC cohort.

### Absolute risks at fixed times

High-pTT groups demonstrated higher event-free proportions at 12 and 24 months. For DMFS, the 12- and 24-month estimates were 85.7% and 77.3% in the high-pTT group versus 75.4% and 65.8% in the low-pTT group, respectively (absolute differences 10.3% and 11.5%). For PFS, the corresponding values were 78.7% and 72.8% versus 68.7% and 56.7% (absolute differences 10.0% and 16.1%), respectively. For OS, 12- and 24-month survival was 94.4% and 85.7% versus 87.0% and 69.7% (absolute differences 7.4% and 16.0%). Consistent with these differences, medians were not reached for the high-pTT groups, whereas low-pTT medians were approximately 40.0 months (DMFS), 30.4 months (PFS), and 50.4 months (OS), respectively.

### Competing risk and cause-specific analyses

Accounting for death prior to distant relapse as a competing event, higher pTT remained associated with a lower subdistribution hazard of distant relapse (Fine–Gray sHR per five percentage-points, 0.968; 95% CI, 0.938–0.998; p=0.038). Cumulative incidence curves separated with borderline significance (Gray’s test p=0.059), whereas the competing event (death before distant relapse) did not differ by pTT (p=0.661) (Supplementary Figure. 4). A cause-specific Cox model yielded a similar association (cause-specific HR = 0.967, 95% CI: 0.937–0.998; p=0.034), with proportional hazards satisfied (global p = 0.83). Event counts were concordant with these findings (distant relapse: 13 in high-pTT vs 58 in low-pTT; competing deaths: 8 vs 26).

### pTT and treatment-related toxicity

#### Pneumonitis or Radiation Pneumonitis

In the MSKCC cohort, clinically significant (grade ≥2) and severe (grade ≥3) pneumonitis occurred more frequently among patients with lower than median pTT values. Specifically, high-grade pneumonitis was observed in 13.9% (n = 16) of patients with low pTT compared with 5.2% (n = 6) among those with high pTT (p = 0.04). This association remained consistent after stratification by volume of the lungs receiving ≥ 20 Gy (lung V20), an established dosimetric predictor of pneumonitis. Among patients with lung V20 above the median, severe pneumonitis (grade ≥3) occurred in 20.8% of those with low pTT versus 9.1% of those with high pTT. Among patients with lung V20 below the median, corresponding rates were 6.8% and 1.9%, respectively. Thus, patients with both high lung V20 and low pTT represented the subgroup at greatest risk (20.8%, n = 11), whereas those with low lung V20 and high pTT had few observed events (1.9%, n=1) (Figure 5A). Overall, the predicted risk of severe pneumonitis decreased with increasing pTT, as modeled using restricted cubic splines with 5-fold cross-validation (Figure 5B)

**Figure 5.**
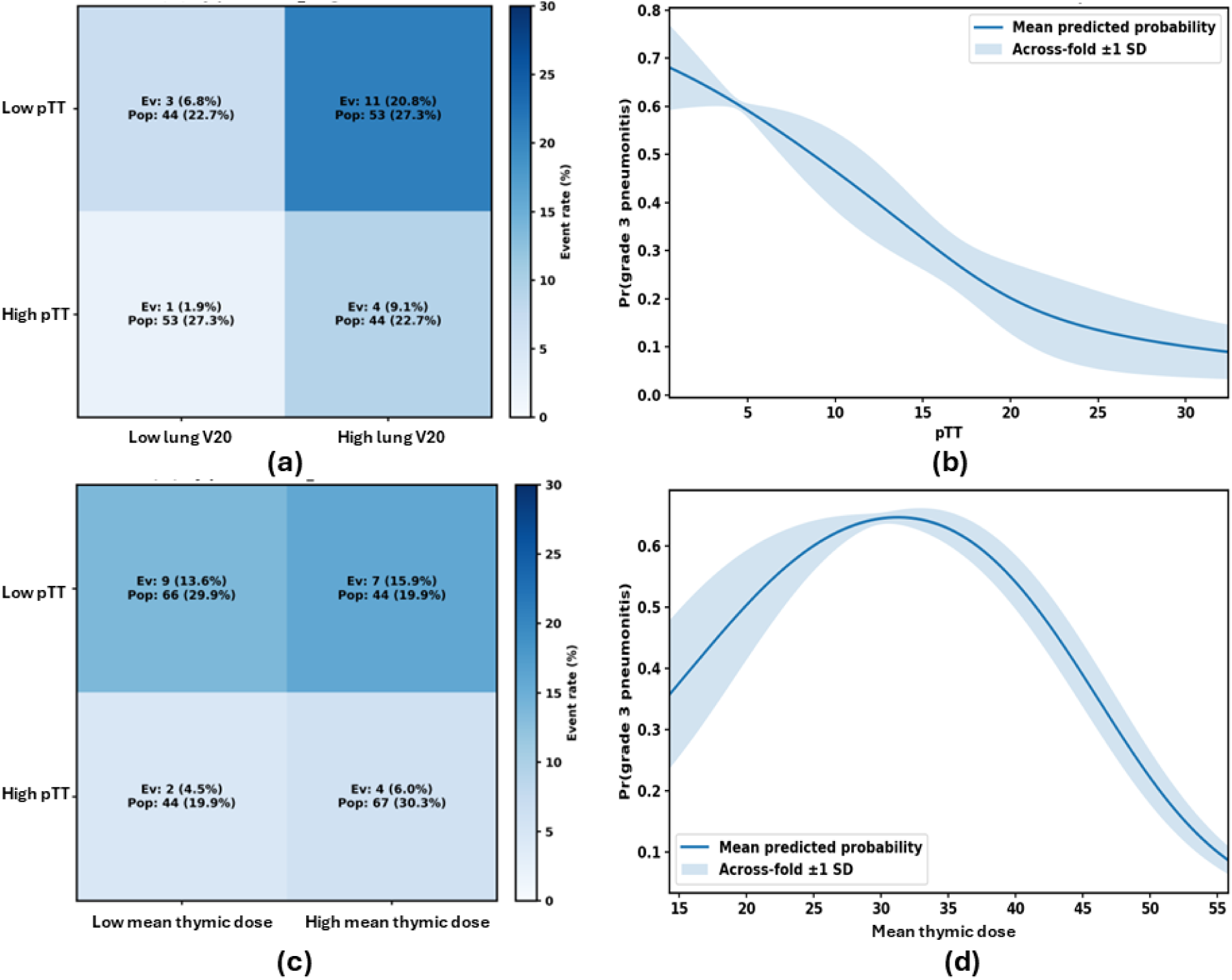
pTT based discrimination of grade 3+ pneumonitis in MSKCC cohort a) pneumonitis events stratified by lung V20 and pTT, b) effect curve for pneumonitis with five-fold cross validation demonstrating decreasing pneumonitis risk with increasing pTT (Fisher p-value: 0.0029). c) pneumonitis events stratified by Mean thymic dose, d) effect curve for pneumonitis with five-fold cross validation demonstrating increasing pneumonitis risk with increasing mean thymic dose (Fisher p-value: 0.015) up to a threshold of 30 Gy.Fisher p values are calculated based on the model using ROC based analysis.

#### Overall immunotherapy tolerance

In the MSKCC cohort, analysis of the association between pTT and clinically significant, treatment emergent AEs leading to durvalumab discontinuation demonstrated that pTT was most predictive among patients with stage IIIC NSCLC with area under a receiver operating characteristic (ROC) curve (AUC) of 0.797. Predictive performance was lower among patients with stage IIIB (AUC = 0.578) and stage IIIA disease (AUC = 0.566), with an overall AUC of 0.55 across all stages (Figure 6).

**Figure 6.**
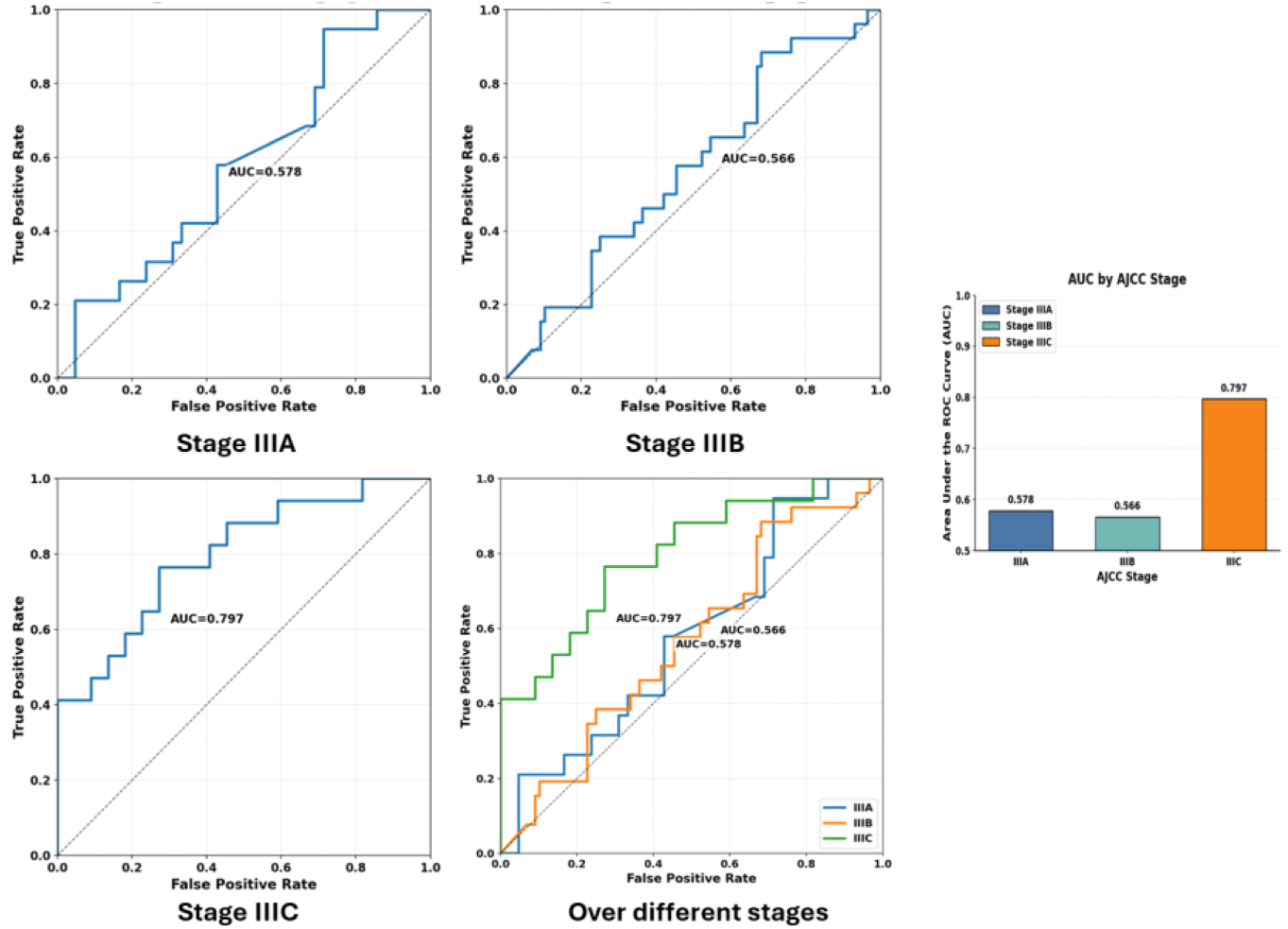
pTT based discrimination of clinically significant toxicity leading to definitive durvalumab discontinuation using Receiver Operating Characteristics (ROC) curves and Area under the ROC Curves (AUC) for stage IIIA, IIIB and IIIC patients.

### Radiation dose sensitivity

The amount of radiation deposited to the thymic region during radiotherapy was independently associated with a higher risk of distant relapse, with volume of the thymus receiving ≥20 Gy (thymus V20) associated with DMFS (aHR 2.01; 95% CI, 1.016–3.995; p = 0.045) (Supplementary Figure 5). However, adjusting for thymus V20 did not attenuate the association between pTT and DMFS (aHR per five percentage-points, 0.968;CI, 0.938–0.998; p = 0.037). As an exploratory negative-control endpoint, pTT was also associated with reduced local failure (aHR per five percentage-points, 0.960; 95% CI, 0.928–0.992; P = 0.014), with the association persisting after adjusting for V20 (aHR, 0.955; 95% CI, 0.924–0.988; p = 0.0075); given the dose/target dependence of local control, this observation is interpreted with caution. Radiation dose to the thymus appeared to be associated with increased risk for severe pneumonitis, independently and in combination with baseline pTT. Patients with both high mean thymus dose and low pTT represented the subgroup at greatest risk for pneumonitis (15.9%, n = 7), whereas those with low mean thymus dose and high pTT had the least risk (4.5%, n = 2) (Figure 5C). Overall, the effect curve for severe pneumonitis with five-fold cross validation demonstrated increasing pneumonitis risk with increasing mean thymic dose up to a dose of 30 Gy (Fisher p-value: 0.015) (Figure 5D).

## Discussion

We developed a robust and interpretable non-invasive quantitative imaging metric for thymic tissue composition, called the percent of thymic tissue, which directly captures age-related glandular atrophy and was independently associated with survival and toxicity outcomes in locally advanced NSCLC. In two independent cohorts totaling 464 patients with baseline CT images obtained using different scanning protocols, we observed consistent associations between thymic tissue composition and other host demographic factors, including sex, with higher baseline thymic tissue proportions in female patients. These findings support the biological plausibility, reproducibility, and generalizability of our proposed thymus composition metric across different populations and settings. Importantly, higher pTT was linked to better survival outcomes in both the MAASTRO discovery cohort and the MSKCC validation cohort, where pTT also effectively stratified risk for local and distant progression as well as treatment-emergent adverse events leading to the discontinuation of durvalumab. These associations remained significant after adjusting for key clinical variables, including age, sex, histology, and smoking status.

The thymus, where T cell progenitors differentiate and undergo selection, is crucial for immune function by producing a diverse range of naïve T lymphocytes, including those with tumor-reactive potential ^41,42^. Mesenchymal stromal cells (MSCs), together with specialized thymic epithelial cells (TECs), contribute to the thymic microenvironment that supports functional T lymphocyte development^43,44^. Although thymic atrophy is a well-described feature of aging, it is not complete, and thymic activity can persist into late adulthood^5,8,35^. The amount of residual thymic tissue varies significantly among individuals of the same age, especially under chronic stressors like smoking, cancer, and cytotoxic treatments^7,22^. Our findings indicate that residual thymic tissue, as readily quantified on routine CT scans, reflects an underrecognized aspect of host immunity and immunosurveillance capacity, influencing cancer outcomes after definitive treatment.

An important potential confounder is radiation deposited to the thymic region during treatment, which may further potentiate lymphopenia by accelerating involution of the gland and reducing the efficacy of thymopoesis^4,18^. Indeed, in the MSKCC cohort, where radiotherapy plans were also available for analysis, higher radiation dose to the thymic region was independently associated with an increased risk of metastatic relapse, presumably due to the resultant impaired tumor immunosurveillance. However, radiation dose did not attenuate the predictive value of pTT. Radiotherapy for thoracic malignancies often results in the delivery of significant doses to the thymic region, potentially inadvertently damaging the organ^18,37,45–47^, but the specific dose that causes treatment-related thymic injury remains unclear. This is partly due to the heterogeneity of the thymic microenvironment, which includes relatively radioresistant MSCs that support T-cell development alongside TECs, and highly radiosensitive lymphocyte populations^48–50^. Preserving MSCs and TECs during treatment may, therefore, help maintain immune resilience, allowing for better recovery of T-cell function during the post-chemoradiation convalescence phase and improving long-term tumor surveillance. The association we found between higher pTT and better survival as well as the impact of radiation dose to the thymic region on distant relapse risk, supports thymic composition as a biologically plausible mechanism through which treatment-related injury may compromise systemic immunity, blunting host capacity to eliminate or suppress disseminated tumor cells in the metastatic cascade^51,52^. This observation emphasizes the importance of thymic structural integrity in adult cancer patients.

Our findings, while hypothesis-generating, have potentially significant implications for thoracic oncology practice across therapeutic modalities. For radiotherapy, ionizing radiation affects lymphocyte counts both directly, through exposure of circulating lymphocytes, and indirectly, via damage to primary and secondary lymphoid organs^17,37,53,54^. Although often considered vestigial, functionally inconsequential and not included among organs-at-risk (OARs) in radiotherapy planning, the thymus may serve as a latent reservoir for T lymphocyte regeneration, especially in patients undergoing such intensive treatments. Our data suggest that preserving thymic tissue integrity, potentially through enacting stringent dose constraints or adaptive radiotherapy that minimize dose to the thymic region, could enhance immune recovery and improve long-term treatment outcomes. We also found that there is a lower incidence of severe pneumonitis among patients receiving a very low dose to the thymic region. This is presumably because sparing the thymus helps maintain regulatory T-cell renewal and immune homeostasis, thereby reducing the risk of unchecked inflammatory reconstitution in lung tissue following chemoradiation^55,56^. Therefore, including thymic dose constraints or protective strategies may warrant investigation in future trials^18,45,57,58^.

Our pTT measure also gains relevance in the immunotherapy era, where ICIs have been integrated in the management of patients with locally advanced or metastatic NSCLC. ICIs and adaptive T-cell therapies depend on effective immune function and clonal diversity, both of which may be influenced by thymic output ^39,57,59–61^. While both the MAASTRO and MSKCC cohorts received chemoradiotherapy, patients in the MSKCC cohort also received the modern standard of consolidation durvalumab, making this the more clinically relevant population for validation and application to current practice^11,62^. Stratifying patients by baseline thymic tissue composition could improve selection for consolidation immunotherapy by identifying those at increased risk of early progression after chemoradiation, predicting pneumonitis-related ineligibility for durvalumab, or revealing a predisposition to immune-mediated AEs^63,64^. Indeed, in our study, we found that among patients with more advanced locoregional disease (stage IIIC), pTT predicted discontinuation of consolidation durvalumab due to treatment-related toxicity. We postulate that this association reflects the heightened antigenic and inflammatory milieu accompanying bulky or multistation N3 disease involvement. Greater tumor burden may elicit amplified immune activation following chemoradiation and PD-(L)1 blockade^65^, particularly in patients with a robust immune system, thereby predisposing these individuals to increased immune-related AEs^66–68^. With regards to the link between low pTT and severe pneumonitis, it is worth noting that after concurrent chemoradiation, the immune system experiences profound perturbation^62,69^. In those with low pTT, there is likely to be impaired recovery of regulatory T cells (Tregs), which normally limit inflammatory responses. Under PD-(L)1 blockade, the effector compartment (CD8+, Th1) may rebound disproportionately during post-chemoradiation convalescence, creating a proinflammatory milieu. The resultant exaggerated, poorly regulated immune reactivation in the lung parenchyma may drive the development of more severe pneumonitis. This aligns with immunosenescence literature showing that thymic involution predisposes to both ineffective anti-tumor immunity and auto-inflammatory toxicity when checkpoints are inhibited^55,70,71^. While there might also be an interaction with radiation dose to the thymic region, the relative immunologic insult is probably greater in patients with low pTT. Overall, the predictive and prognostic value of pTT across different treatment settings and endpoints, albeit at different thresholds, likely reflects its utility across various therapeutic strategies^11,16^. Moreover, longitudinal monitoring of pTT after treatment might also provide insights into immune reconstitution and late toxicity, particularly in survivorship following curative therapies.

The proposed modeling framework addresses several limitations of prior imaging-based approaches. Existing methods, such as those introduced by Okamura et al ^24^, relied on kernel density estimation (KDE) to describe attenuation profiles and excluded patients with nonstandard CT histograms. This exclusion introduces survivorship bias and limits generalizability, as mixed tissue compositions are common in aging and diseased glands ^21,22^. In contrast, we utilized a parametric model, namely Gaussian Mixture Model (GMM) to decompose Hounsfield Unit (HU) distributions into component tissue classes, enabling inclusion of heterogeneous or ambiguous attenuation patterns. Additionally, GMM model improves interpretability as compared to the non-parametric KDE-based model used by Okamura et al.^24^. The GMM-based estimation of mean attenuation avoids fixed thresholds and provides a probabilistic framework for assigning tissue class membership^32,33,72^. Our pTT metric is volume-independent, segmentation-aware, and interpretable as the proportion of thymic tissue relative to adipose infiltration, microvessels, inflammatory changes, or glandular scar elements. This formulation allows meaningful comparisons across patients, regardless of body size, scan protocol, or gland morphology. In contrast to morphometric surrogates or deep learning-derived features, pTT remains grounded in biologically interpretable units, supporting its use as a generalizable quantitative imaging biomarker ^25,73^. This is especially important in translational settings where mechanistic insight is essential. Although previous studies have shown that mean HU correlate with age, such correlations are insufficient to capture the immunologic effects of structural loss^21–23^. Our analysis demonstrates that pTT not only reflects age and sex differences but also provides predictive and prognostic information that is independent of these factors. Furthermore, pTT consistently outperformed chronological age alone in predicting survival, suggesting that thymic tissue composition offers orthogonal insight into physiological reserve not captured by demographic variables.

To our knowledge, this is the first study to analyze the relationship between a directly interpretable and readily generalizable radiographic marker of thymic composition, cancer outcomes,and treatment related toxicity. Okamura et al, introduced a method to quantify thymic involution and hyperplasia using global kernel density estimation on a small sample of subjects with CT scans^24^. Our extensive multi-cohort study employs more detailed statistical modeling that captures distinct components of the thymic microenvironment in a cancer population. Conversely, while deep learning can extract complex features, these “black-box” methods are often prone to bias arising from human annotated training data, underepresentation of diverse populations in trainings sets, and lack of interpretability; factors that collectively limit their generalizability, translational potential, and overall clinical utility^30,39,73,74^. Gaussian mixture modeling, by contrast, is a probabilistic, unsupervised method with clear assumptions and outputs that can be easily interpreted and understood without biased pre-training, and was therefore the preferred method for our study. Our pTT metric is also rooted in biological plausibility, quantifying thymic tissue proportion in a reproducible and clinically meaningful way that does not obscure how imaging features translate into indices and models heterogeneity across all patients continuously^73^. It captures substructures and partial function even in the setting of damage to the organ, without introducing artificial discontinuities into the data by hand-selecting cases for study. pTT is directly applied to all imaging data, avoiding reliance on pre-defined HU thresholds or pre-trained models or non-representive external training datasets, and can be used as a reliable, biologically grounded standard for studying the thymus using radiographic data without embedded biases.

Our study has limitations, including its retrospective design. Additionally, in the MAASTRO dataset, patients were treated with an older chemoradiotherapy approach, and OS was the only available clinical endpoint; OS is heavily confounded and may not be as meaningful or actionable in a clinical context, especially for patients receiving immunotherapy. These limitations were mitigated by validation in the MSKCC cohort, which received the current standard of care with consolidation durvalumab^11,62^ and included multiple relevant and actionable outcomes such as DMFS, LRF, PFS, and OS as well as toxicity data. Our analysis, however, depends on imaging-based metrics that are indirect surrogates of underlying thymic composition and output. Further research integrating T-cell receptor sequencing, measures of thymic emigrants, or functional immune assays could provide direct evidence of thymic activity and could clarify the biological importance of structural preservation ^6,35,39,75–78^. We also focused exclusively on the thymus, although the broader immune system includes contributions from the bone marrow, spleen, and secondary lymphoid organs^1,2,79^. Future studies including radiomics and/or volumetric analyses of these compartments to construct a more comprehensive model of systemic immune capacity are warranted. We are also currently working on generating a spatially aware pTT measure that identifies specific areas within the contoured thymic region likely to harbor the most residual thymic tissue, and thus enabling them to be prioritized in treatment planning. A measure that captures radiation dose to the volume of residual thymic tissue (DpTT) can conceivably be generated in this precision-guided radiotherapy paradigm.

In conclusion, pTT offers an interpretable, readily generalizable, scalable, and prognostically valuable measure of thymic integrity in adult lung cancer patients. This framework unites statistical rigor with biological interpretability and broad clinical applicability. By providing a novel lens into the concept of structural immune reserve, pTT may serve as a foundation for future efforts to personalize treatment based on an individual’s intrinsic capacity for immune recovery and to better characterize the thymus as an increasingly recognized organ-at-risk in lung cancer therapy. Further investigation is warranted to validate pTT as a predictive indicator of outcomes and clinically significant toxicity and as a tool to optimize the balance between efficacy and safety in multimodality lung cancer treatment.

## Methods

### Study population

A total of 464 patients with Stage III NSCLC from two distinct cohorts were included in our analysis. The first cohort, MAASTRO (*n* = 277)^40^, comprised patients who received radiotherapy for locally advanced (stage III) NSCLC at the MAASTRO Clinic in the Netherlands. The second MSKCC cohort (*n=187*) included patients who were treated with chemoradiotherapy followed by adjuvant durvalumab at MSKCC. We used a deep learning pipeline for segmenting the thymic region from contrast-enhanced axial chest CT images. We then developed a novel region-based radiographic parameter, percent thymic tissue (pTT), to quantify thymic composition within the Thymic Region for Quantification (TRQ). The derivation of our pTT metrics is described below (Figure 1).

### Derivation of percent thymic tissue (pTT)

#### Thymic region definition

A thymus contouring atlas was defined in consultation with a board-certified thoracic radiologist at MSKCC. The thymic region was contoured according to consistent anatomic landmarks on axial CT imaging. The superior border was defined at the level where the left brachiocephalic vein crosses midline, which is often accompanied by a small fatty space posterior to the sternum. The posterior surface of the sternum marked the anterior border, while the posterior border was delineated by the adjacent great vessels. The mediastinal pleural surfaces defined the lateral borders. The inferior border was set at the level of the bottom of the arch of the right pulmonary artery, which corresponds to the uppermost axial slice of the automatically generated heart contour used for radiotherapy planning at our center (Thymus Contouring Atlas in the Supplements; Supplementary Figure.1).

#### Thymic region contouring

Manual thymic region contours were generated on axial chest CT images by a board-certified thoracic radiation oncologist and a senior radiation oncology resident at MSKCC using 3D Slicer v5.4.0^80^ and MIM Software v7.3.7 (MIM Software Inc., Cleveland, OH, USA). A deep learning-based framework based on Swin Transformer was then utilized to automate the contouring process with human-in-the-loop quality control. At least two other thymus autosegmentation models on chest CT based on convolutional neural-networks have been published ^24,34,38^; however, for our pipeline, we opted to employ a Swin transformer based deep learning model, termed, SWIMMER, to automatically define the *Thymic Region for Quantification* (TRQ; technical convention retained for consistency with Okamura et al.), which was used for all downstream analyses. Detailed descriptions of model architecture and implementation for the thymus and other thoracic and lymphoid structures, will be reported in a separate technical manuscript (a concise summary is provided in the Supplementary Methods). In the supplemental material, we also highlight key aspects of the framework and situate SWIMMER relative to existing open-source thymus autosegmentation approaches. Training used expert voxel-wise annotations on volumetric CT with deep supervision, postprocessing applied 3D connected-component filtering to remove discontinuous false positives and retain anatomically plausible anterior-mediastinal structures. The resulting mask defined the thymic region or TRQ, which served as the reference for all volumetric, spatial, and radiographic composition analyses.

#### HU histogram extraction

For each patient, a histogram of Hounsfield Unit (HU) values was computed from the Thymic Region for Quantification (TRQ) by extracting all voxel intensities within the segmentation mask. Only finite HU values were retained, without applying thresholding, windowing, or outlier removal. The HU values were binned into 50 equal-width intervals spanning the full dynamic range observed in the TRQ (approximately from –150 to +100 HU). The histogram was then normalized to reflect the empirical probability distribution of attenuation values within the segmented thymus, thereby capturing the composite density of residual thymic tissue, infiltrating fat, and edge voxels with ambiguous radiodensity. These may result from partial volume effects or inclusion of adjacent mediastinal structures due to minor segmentation imprecision.

#### Gaussian mixture modeling of HU distributions

To decompose the heterogeneous HU distribution into physiologically meaningful tissue components, we modeled the TRQ histogram as a weighted sum of k Gaussian densities:

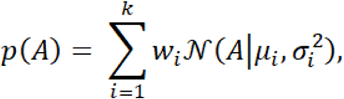

where A denotes the attenuation (HU), w_i_, the weight of *i*^*th*^ gaussian component, and N being the normal distribution with mean μ_i_ and variance σ_i_^2^. The parameters of the gaussian mixture model have been estimated using the Expectation-Maximization (EM) algorithm^72^. The optimal number of components k was selected by minimizing the Bayesian Information Criterion (BIC), allowing model complexity to adapt to anatomical heterogeneity.

We considered both non-parametric and parametric methods for estimating HU density. While kernel density estimation (KDE) provides a smooth, non-parametric estimate of this distribution, it lacks interpretability in terms of discrete tissue classes and the selection of a single mode value might obscure other biologically meaningful modes. In contrast, Gaussian mixture modeling (GMM) offers a principled framework to model the histogram as a weighted sum of underlying tissue components, enabling reproducible decomposition into adipose- and thymus-dominant compartments with interpretable parameters. Compared to non-parametric approaches such as KDE, which rely on fixed bandwidth smoothing and lacks component interpretability, GMMs offer a flexible yet principled means of modeling multimodal distributions. Each Gaussian component corresponds to a biologically interpretable tissue class (e.g., adipose, thymic tissue), and the model permits likelihood-based selection criteria to determine the number of meaningful classes. KDE, by contrast, may over-smooth or obscure biologically distinct modes, particularly in lower-volume or high-variance distributions. Thus, GMMs were better suited to isolate tissue compartments in a reproducible and quantifiable manner. GMM being a parametric approach and being a universal approximator for smooth densities^82^, we considered GMM based analysis in the sequel.

The mean HU of the TRQ, denoted A_TRQ_, was computed as the weighted sum average of individual Gaussian means:

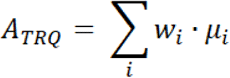

To minimize false low estimates due to segmentation error or volume averaging with perithymic fat, a Bayesian correction was applied:

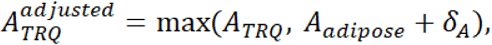

where A_adipose_ is a canonical adipose attenuation value (typically −100 HU) and δ_A_ is a small offset (1 HU) to avoid spurious misclassification of fat-only regions.

#### Estimated thymic volume (ETV) and percent thymic tissue (pTT)

Under a two-compartment model in which the TRQ comprises thymic tissue and adipose tissue with characteristic attenuations A_thymic_ and A_adipose_, the TRQ can be represented as a density-weighted mixture rule:

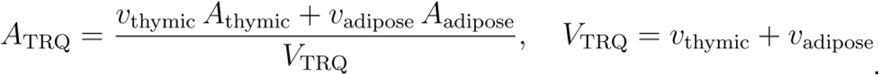

Solving for the thymic volume yields

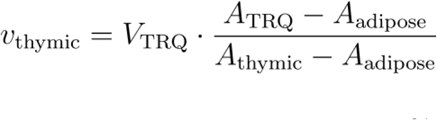

In their prior work, Okamura et al.^24^ defines the TRQ by summarizing a single HU value, *A*^*^_TRQ_ given by the KDE-estimated mode of the HU distribution, and mapped to a hard thymic fraction using fixed anchors (*A*_adipose_, *A*_thymic_); the resulting estimator of thymic content is

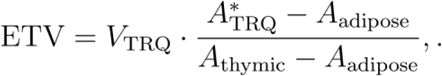

By construction, ETV is volume-dependent and relies on a single-value summary of the HU distribution. Because of this, ETV scales linearly with the size of the segmented TRQ; its utility for between-subject comparison is limited regardless of modeled tissue heterogeneity, necessitating normalization for volume to isolate composition. Building on this framework, we replace the single-value fraction with a distribution-aware fraction derived from the GMM. To derive a volume-invariant index, we defined percent Thymic Tissue (pTT) by letting *γ*_*vi*_ = *P* (*Z* = *i* | *A_v_*) denote the posterior responsibility that voxel *v* belongs to mixture component *i*, and letting ℱ denote the adipose component set (the lowest-mean mode optionally including any sub-fat mode). We then specify a generalized estimator *ETV*_GMM_ as the TRQ volume multiplied by the posterior-averaged non-adipose fraction, and define our primary composition index as

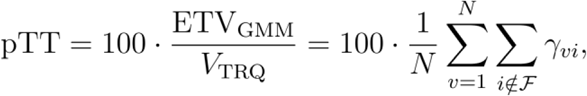

which is volume-invariant by construction and leverages the full attenuation distribution while down-weighting partial-volume and ambiguous voxels via soft assignments. In the unimodal, anchor-consistent limit assumed by Okamura et al.^24^, this reduces to pTT ≈ 100 · ETV/*V*_TRQ_.

This formulation ensures that pTT remains invariant under TRQ volume scaling, making it more suitable for intersubject and longitudinal comparison. pTT thus provides a normalized, biologically interpretable index of thymic composition that accounts for both tissue attenuation and segmentation variability.

#### Clinical Outcomes

The analyzed outcomes, where available, were (1) distant metastasis free survival (DMFS), defined as the interval from the start of radiotherapy to the occurrence of distant metastasis, measured in months, (2) locoregional failure (LRF), defined as the interval from the start of radiotherapy to the occurrence of local or regional recurrence, measured in months; (3) progression free survival (PFS) defined as the interval from the start of radiotherapy to distant or locoregional progression, measured in months, and (3) overall survival (OS), defined as the time from the start of radiotherapy to death from any cause, also measured in months. DMFS, LRF, PFS, and OS data were collected for MSKCC patients, and only OS was available for the MAASTRO cohort. In addition to time-to-event analyses, absolute event-free proportions at fixed times (12 and 24 months) estimated by the Kaplan–Meier method were key secondary end points.

#### Statistical analysis

Standard statistical measures were used to characterize clinical and demographic features, thymic composition parameters, and outcome measures. Group comparisons were conducted with Wilcoxon rank-sum test for continuous variables and regression analyses as appropriate. Time-to-event outcomes (including standard clinical endpoints of DMFS, LRF, PFS, and OS) were estimated using the Kaplan-Meier methods, with hazard ratios derived from proportional hazard models in both univariable and multivariable settings.

We evaluated the prognostic value of pTT in two independent cohorts of patients with NSCLC. We studied the associations between calculated pTT and OS, LRF, PFS, and DMFS, using Cox proportional hazards models adjusted for relevant clinical covariates and Kaplan-Meier analysis. Hazard ratios and their statistical significance were reported for each variable in both univariate and multivariable contexts. Optimal threshold for pTT were identified using maximally selected rank statistics and were optimized for each survival endpoint, as is standard. Wilcoxon rank-sum test and linear regression were used to assess the differences in pTT across sex and age. pTT was analyzed both as a continuous variable and dichotomized at the cohort-derived optimal thresholds. For continuous modeling, effects were scaled per 5 percentage points of pTT, and potential non-linearity was assessed using restricted cubic splines. For DMFS, death before distant relapse was treated as a competing risk; subdistribution hazards were estimated using Fine–Gray models with cumulative incidence compared by Gray’s test, and cause-specific Cox models were fit as a sensitivity analysis. In the MSKCC cohort, radiation dose–response was evaluated by incorporating thymus V20 (the relative TRQ volume receiving ≥20 Gy) extracted from DICOM-RT dose mapped to the planning CT; V20 was entered as a continuous covariate in multivariable models. Absolute risks at 12 and 24 months were summarized from Kaplan–Meier estimates (or cumulative incidence for DMFS), and restricted mean survival time (RMST) differences between high- and low-pTT groups were estimated at τ = 24 and τ = 36 months. Model performance was quantified by Harrell’s c-index; time-dependent AUCs at 12 and 24 months and IPCW Brier scores were computed, and calibration at 12 and 24 months was assessed with internal resampling.

All tests were two-sided, with P <0.05 considered statistically significant and 95% confidence intervals were reported. All the analyses were performed in Python version 3.9.21 and R version 4.43.

#### Clinically significant toxicity prediction using pTT

We performed stage-specific analyses to evaluate the association between pTT and clinically meaningful toxicity in patients treated within the MSKCC cohort. Clinically meaningful toxicity was defined as any adverse event resulting in definitive durvalumab discontinuation during a patient’s treatment course. To capture potential non-linear effects of pTT on toxicity risk, we fit third-order polynomial logistic regression models separately for each AJCC stage subgroup (IIIA, IIIB, and IIIC). Additionally, logistic regression with restricted cubic splines (RCS) model was utilized to model the potentially non-linear relationship between pTT, radiation dose to the lungs or thymus, and pneumonitis events. Inverse logit function transformation was applied to the fitted RCS model to obtain the predicted risk probabilities across each predictor. Model discrimination was assessed using Receiver Operating Characteristic (ROC) curves and corresponding Areas Under the Curve (AUCs).

## Ethical Approval

This study was performed in accordance with MSKCC IRB protocol number 16-142 for the retrospective review of thoracic malignancies treated with radiotherapy at the center. It was completed in compliance with institutional research guidelines.

## Acknowledgements

This study was supported in part by the National Cancer Institute Cancer Center Core Grant (P30 CA008748).

## Competing Interests

None

## Data Availability

The study was conducted using an MSKCC institutional cohort of radiotherapy patients obtained with IRB approval, and a publicly available dataset, available for all to download on the TCIA: https://www.cancerimagingarchive.net/collection/nsclc-radiomics/

## Code Availability

Source code for the pTT computation pipeline is available upon reasonable request to the corresponding author and will be openly released following peer review.

## Supplementary Information

### Thymus Contouring Atlas

**Supplementary Figure. 1.**
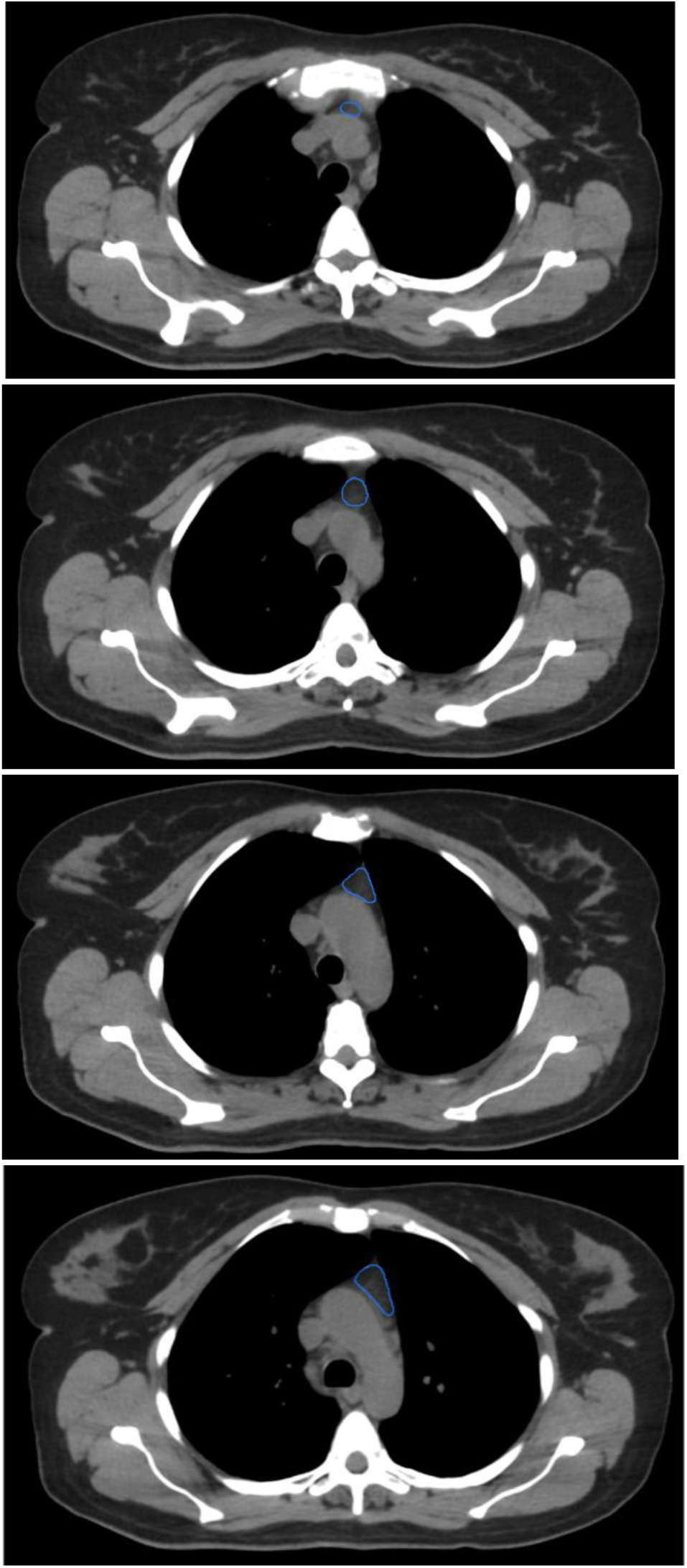
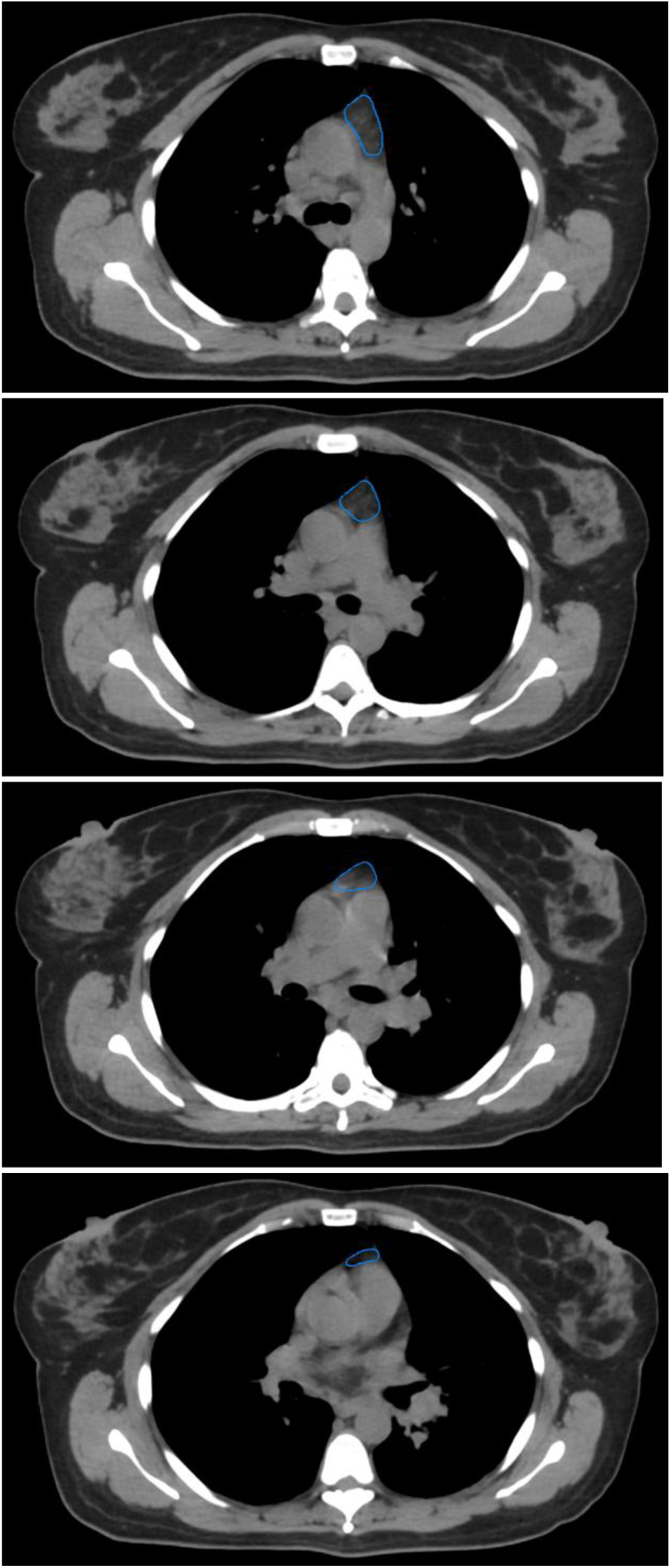
The thymic region was defined using the following borders as demonstrated in this example case with the following standardized anatomical borders; *Superior border:* where the left brachiocephalic vein crosses midline and (sometimes a fatty space appears posterior to the sternum). *Anterior border:* sternum. *Posterior border:* great vessels. *Lateral borders:* mediastinal aspect of pleura. *Inferior border:* bottom of the arch of the right pulmonary artery (superior slice of MSKCC radiotherapy heart contour)

### Segmentation Framework

We developed a modular deep learning pipeline for segmenting the thymic region from contrast-enhanced axial chest CT images using a novel Swin-Transformer based architecture termed SWIMMER (**Sw**in-based **I**mmune & **M**alignant structure **M**apping with **E**ntropy-based **R**egression). The model was implemented in PyTorch using MONAI^36^ and combines the segmentation framework with Monte Carlo (MC) dropout for voxelwise uncertainty estimation. The encoder integrates a Swin Transformer branch for global context and a residual convolutional branch for local detail. The network accepts single-channel CT input and produces the thymus segmentation mask. MC dropout utilized within the framework enabled voxel-wise uncertainty estimation.

The model was trained on expert-annotated scans uses Dice-based losses optimized with AdamW optimizer. Prior to training, DICOM series were converted to NifTI. Uncertainty quantification, when performed, used MC inference with T = 30 stochastic passes. Postprocessing involved 3D connected-component filtering to exclude small disconnected regions or misclassified mediastinal fat; the largest anatomically plausible anterior-mediastinal component was retained, and mask geometry (centroid, volume) was recorded in physical space via the NIfTI affine. All segmentations were manually reviewed for anatomical plausibility; scans with severe artifacts or incomplete mediastinal coverage were excluded. The resulting masks defined the Thymic Region for Quantification (TRQ) and served as the basis for subsequent analysis.

Two published thymus autocontouring models demonstrated feasibility but differ materially from our approach. Okamura et al. introduced the TRQ concept and trained a segmentation network based on DeepLabV3-ResNet-50 to support their ETV metric, however, the downstream pipeline ultimately compresses attenuation to a single HU mode (KDE) and does not expose uncertainty-aware outputs^24,38^. Guo et al. introduced Thy-UNet, an end-to-end deep learning framework that outperformed conventional 3D U-Net architectures as well as more advanced variants, including UNetR, TransUNet3D, and the standard nnU-Net. Thy-UNet was validated on multicenter datasets, including the publicly available radiotherapy cohort used in the present study (MAASTRO)^34,40^. However, this model still relies on a purely convolutional backbone with a limited global receptive field and likewise no voxel-wise predictive uncertainty^34^. A SCA-UNet variant targets *thymic lesion* segmentation on preoperative CT, which is useful for focal pathology, but is optimized for lesion masks rather than reliable whole-gland TRQ delineation in the involuted adult thymus^81^.

SWIMMER advances beyond these methods by combining a Swin Transformer based segmentation model with Monte Carlo dropout for uncertainty quantification. Monte Carlo dropout yields voxel-wise predictive uncertainty that we used for principled QC gating (flagging rare low-confidence masks) without atlas constraints or hand-crafted priors, reducing exclusions from segmentation failure and stabilizing downstream composition analysis (pTT).

## Comparative formulation of ETV versus pTT

Okamura et al.^24^ summarize the thymic region for quantification (TRQ) on chest CT with a single, hard attenuation surrogate, the mode of the TRQ HU density estimated by kernel density estimation (KDE), then assume a two-compartment linear mixture between canonical adipose and thymic attenuations to recover a hard thymic fraction 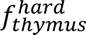 and a volume-scaled estimator of thymic content 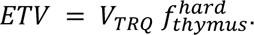 Formally, if 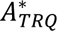 denotes the KDE-mode HU within the TRQ and *A*_*adipose*_, *A*_*thymus*_ are fixed anchors, then

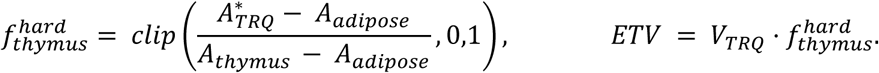

Because *A*^*^_TRQ_ compresses a potentially heterogeneous HU distribution into a single value and ETV scales linearly with *V*_TRQ_, the estimator is both histogram-shape sensitive, with multimodal TRQs often excluded, and volume dependent. Edge voxels, partial-volume effects, perithymic adipose spill-in, scarring, or mixed tissues can therefore perturb the estimate. In contrast, our percent thymic tissue (pTT) treats TRQ HU values 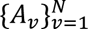 as a distribution and performs Gaussian-mixture decomposition with *K* components to obtain voxel-wise posterior responsibilities (probabilities) *γ*_*vi*_ = *P* (*Z* = *i* | *A_v_*). For clarity, we aggregate thymic (non-adipose) components at each voxel into a single posterior, 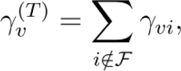 and define a scale-free composition index

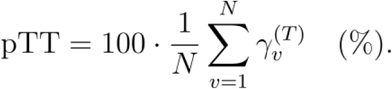

When voxels are anisotropic (or otherwise vary in physical size), we use the volume-weighted analogue:

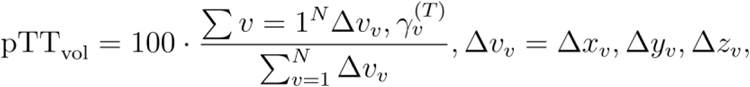

which reduces to the unweighted mean above when voxel sizes are equal (isotropic sampling). This retains heterogeneous cases and down-weights ambiguous, partial-volume, or fibrosis-affected voxels instead of forcing a hard assignment. These design differences manifest empirically in our data. ETV versus TRQ volume shows the expected scaling with a Spearman ρ=0.24, p=6.22×10−7, whereas pTT versus TRQ volume shows a strong biologic inverse relationship(Spearman ρ=−0.67, p=3.77×10−5), consistent with larger adult TRQs being more adipose-replaced (Supplementary Fig. S;). To compare intrinsic composition independent of segmentation size, we removed nonlinear effects of *V*_*TRQ*_ from each metric using a smooth LOESS and GAM (generalized additive model) adjustment and plotted residual versus residual using a spline transform. The residuals align tightly (Pearson r=0.73; Spearman ρ=0.91), confirming both metrics target the same latent biology, yet pTT residuals cluster more tightly and trace a clearer monotone trend than ETV (Supplementary Fig. S;). Collectively, the probabilistic formulation 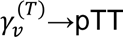 delivers a volume-invariant, heterogeneity-aware, and empirically more stable measure of thymic composition than the hard, volume-scaled ETV, making pTT better suited for cross-patient comparison and downstream prognostic modeling.

**Supplementary Figure. 2.**
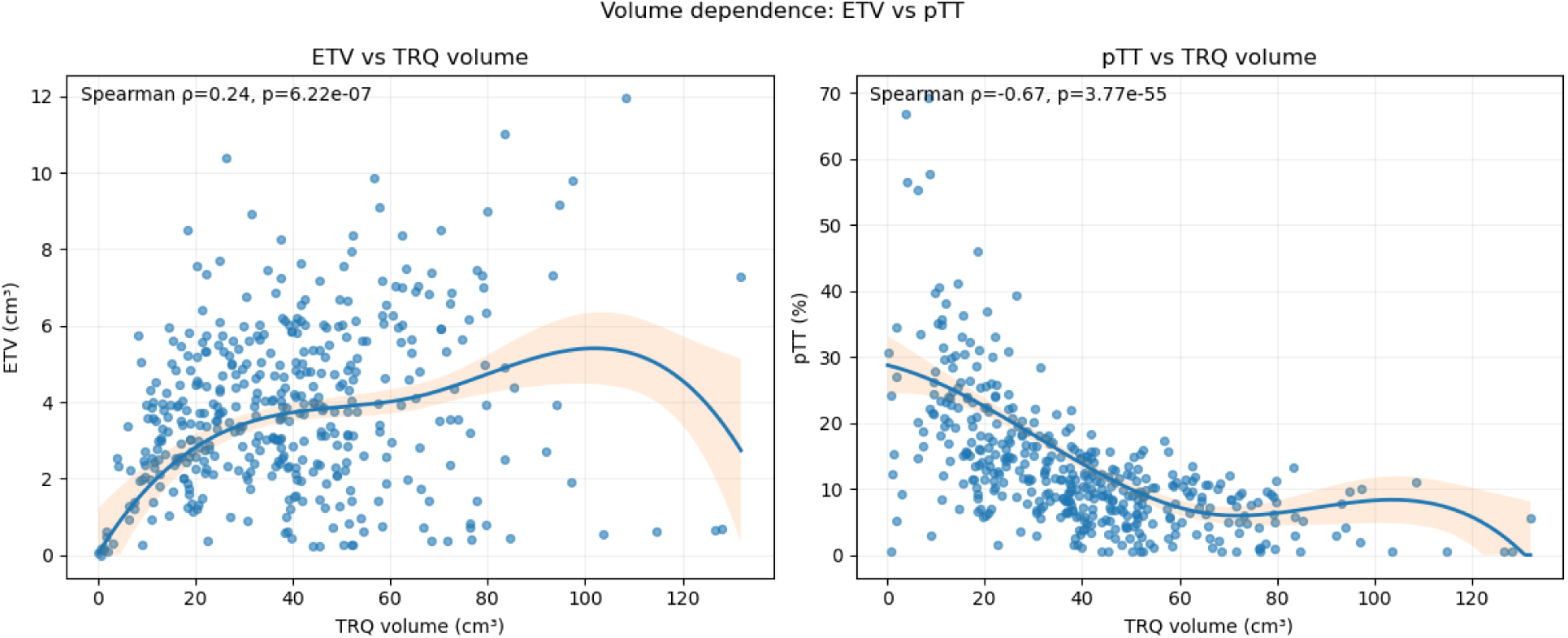
ETV rises with TRQ volume (Spearman ρ = 0.24, P = 6.22 × 10⁻⁷), whereas pTT falls (ρ = −0.67, P = 3.77 × 10⁻⁵⁵), consistent with greater adipose replacement in larger adult TRQs. Curves show LOESS fits with 95% CIs; points are patients (n = 464). Abbreviations: TRQ, thymic region for quantification; ETV, estimator of thymic volume; pTT, percent thymic tissue.

**Supplementary Figure. 3.**
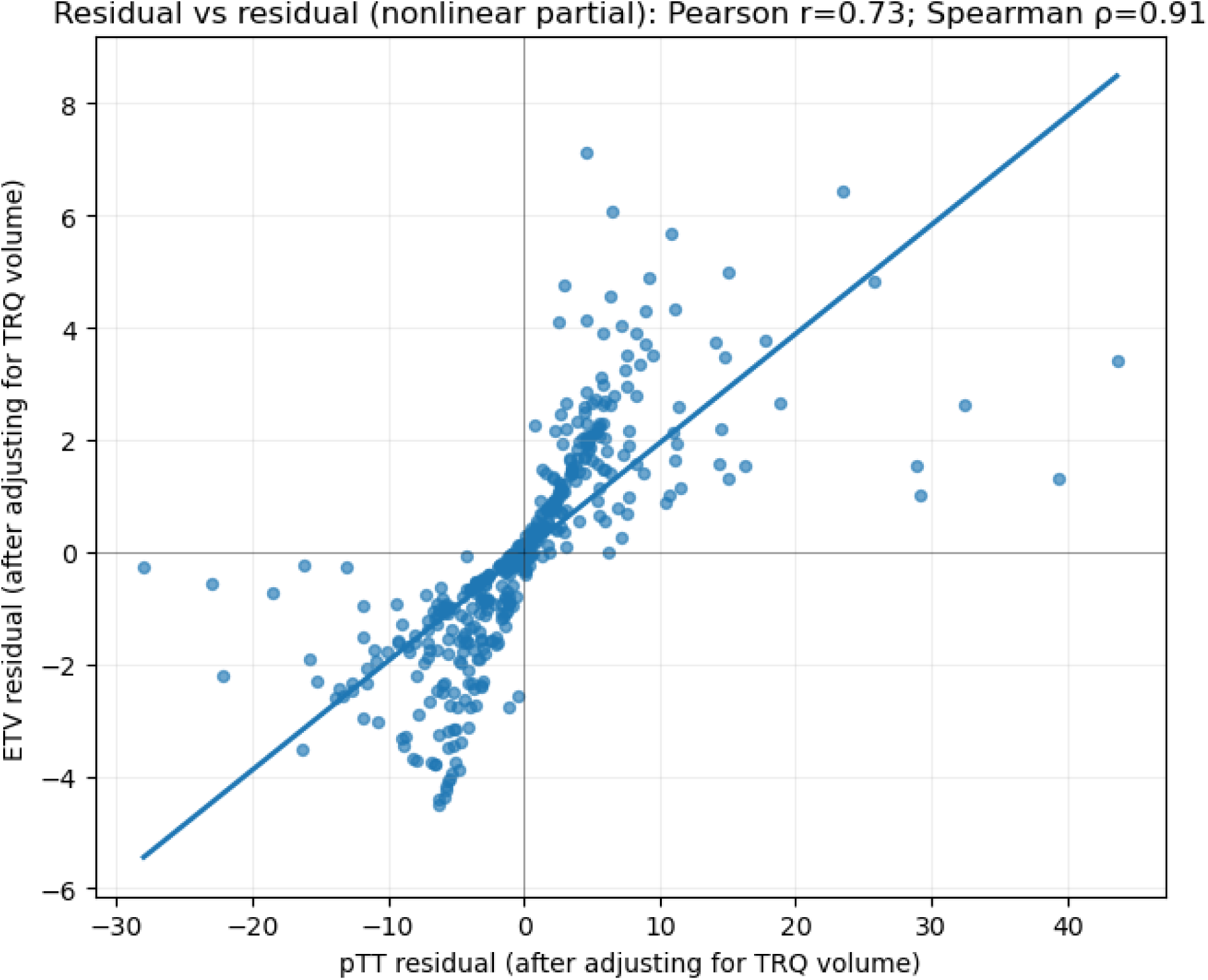
After adjusting both metrics for TRQ volume, ETV and pTT residuals align closely (Pearson r = 0.73; Spearman ρ = 0.91). The tighter dispersion for pTT indicates greater stability to heterogeneity and partial volume. Abbreviations: TRQ, thymic region for quantification; ETV, estimator of thymic volume; pTT, percent thymic tissue.

### Supplemental Results

#### Continuous modeling of pTT

When modeled as a continuous variable scaled per five percentage-points, higher pTT was associated with improved outcomes across endpoints; DMFS (aHR = 0.968, 95% CI: 0.938–0.999, p=0.040), PFS (aHR = 0.973, 95% CI: 0.946–1.002, p=0.064), and OS (aHR = 0.971, 95% CI: 0.943–1.000, p=0.050). Proportional hazards assumptions were met on Schoenfeld testing (global p = 0.87 for DMFS, 0.72 for PFS, and 0.61 for OS).

#### Model performance

For DMFS, adding pTT to a clinical model (age, sex, histology) increased the concordance index from 0.574 to 0.594 (Δ = 0.020; 95% CI, −0.009 to 0.085). Time-dependent AUCs were 0.57 at 12 months and 0.59 at 24 months. Internal resampling showed acceptable calibration at 12 and 24 months. The IPCW-Brier score decreased from 0.207 to 0.141 at 12 months (−0.066 absolute; −31.9% relative) and from 0.243 to 0.192 at 24 months (−0.051 absolute; −21.0% relative) compared with the clinical model alone.

#### Restricted mean survival time

High pTT conferred absolute gains in event-free time. For DMFS, the restricted mean survival time (RMST) difference (high minus low) was +2.0 months at 24 months (95% CI, −0.08 to +4.05; P = 0.060) and +3.6 months at 36 months (95% CI, −0.03 to +7.25; P = 0.052). For PFS, the 24-month RMST difference was +1.7 months (95% CI, −0.70 to +4.11; P = 0.166). For OS, the 24-month RMST difference was +1.8 months (95% CI, +0.30 to +3.27; P = 0.019).

**Supplementary Figure. 4.**
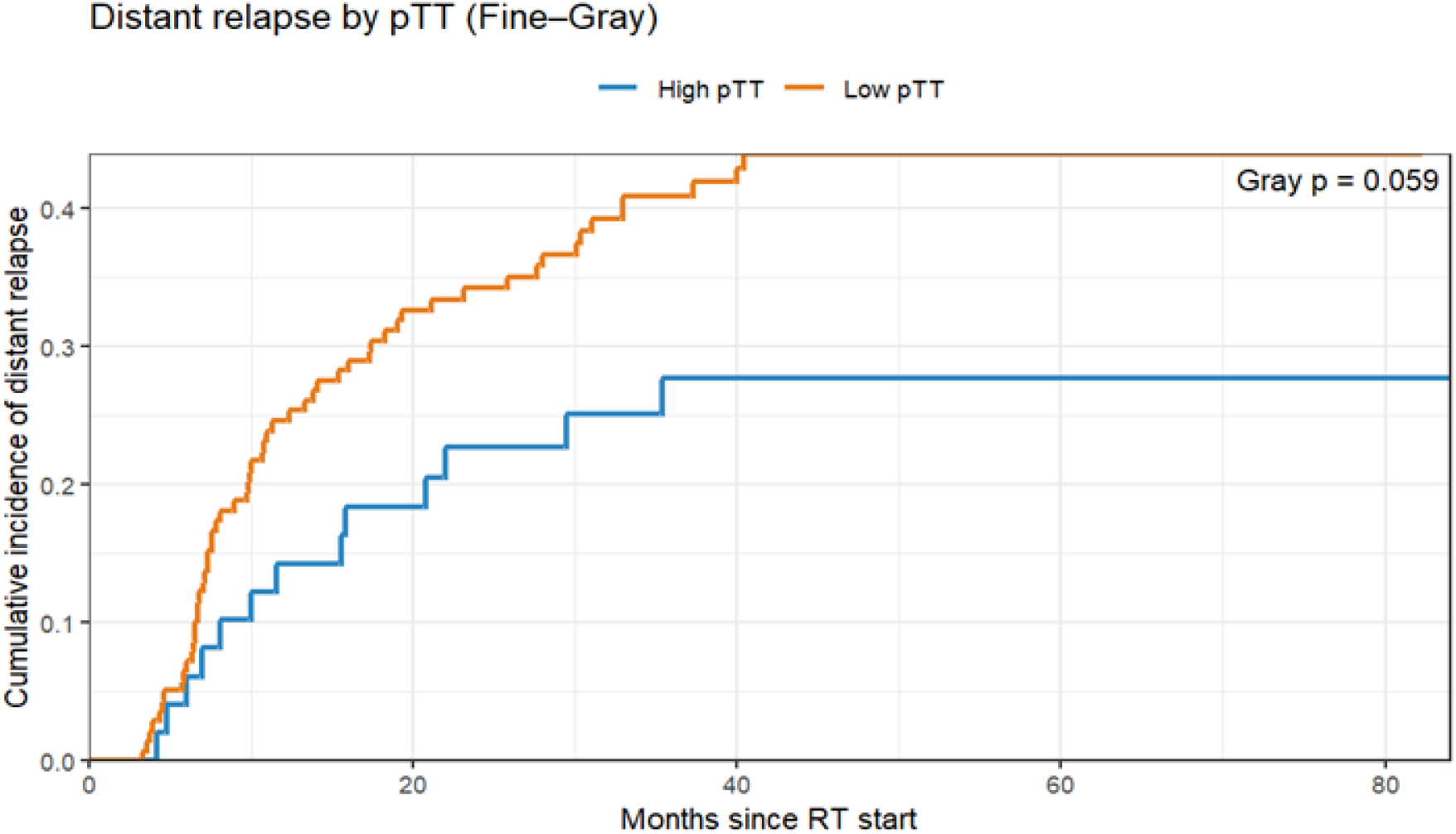
Cumulative incidence of distant relapse by baseline pTT (MSKCC cohort). Fine–Gray competing-risks curves from the start of radiotherapy, with death prior to distant relapse treated as the competing event. Patients are stratified into High pTT vs Low pTT using the cohort-derived cut-point. Lower pTT is associated with a higher cumulative incidence of distant relapse; Gray’s test p = 0.059 (borderline). Abbreviations: pTT, percent thymic tissue; RT, radiotherapy.

**Supplementary Figure. 5.**
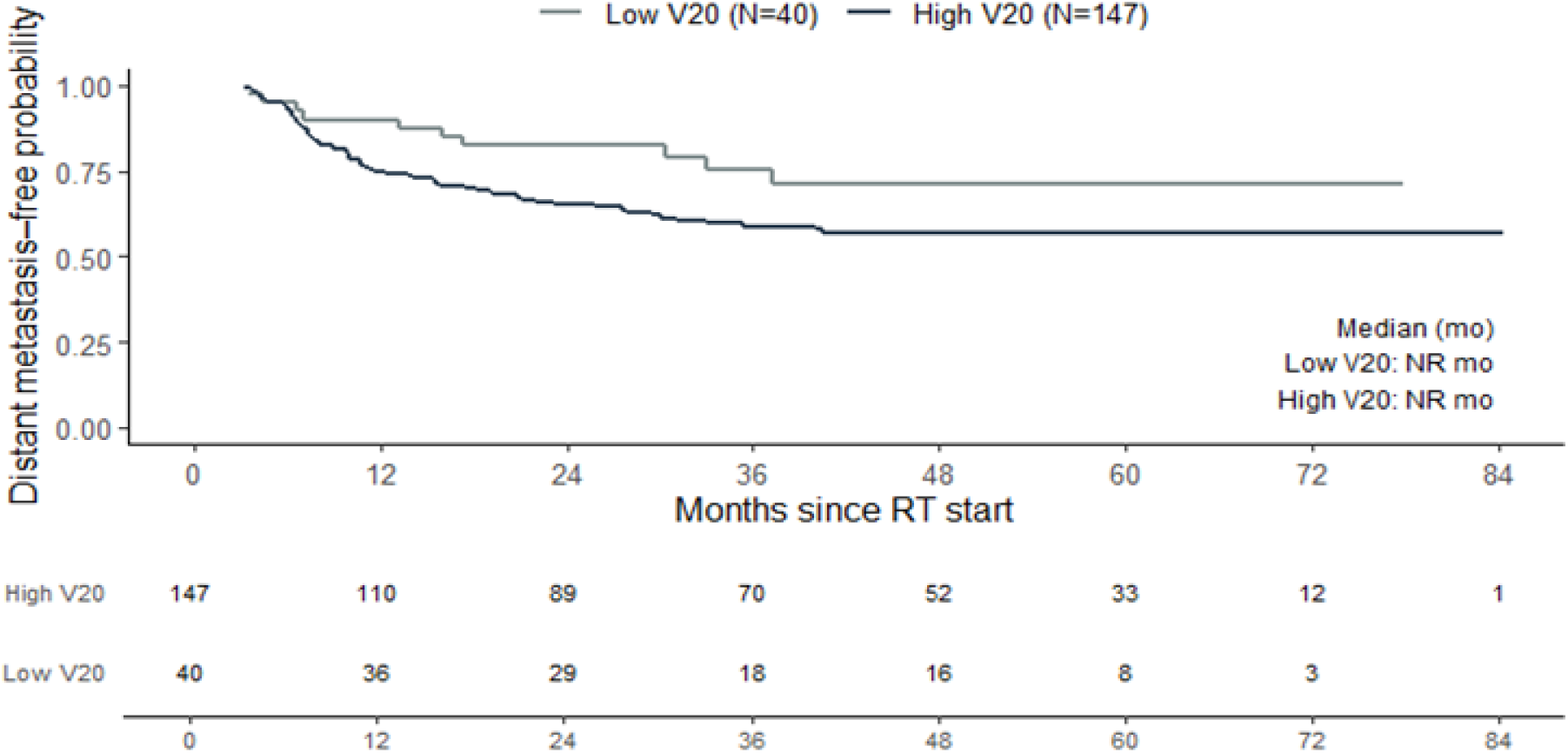
Distant metastasis-free survival (DMFS) by thymus V20 (MSKCC cohort). Kaplan–Meier curves from the start of radiotherapy stratified by thymus V20 (proportion of the TRQ receiving ≥20 Gy on the planning dose distribution). Numbers at risk are shown below the x-axis; medians were not reached (NR) in either group. Higher thymic dose shows a trend toward worse DMFS. Abbreviations: TRQ, thymic region for quantification; V20, % TRQ volume receiving ≥20 Gy; DMFS, distant metastasis-free survival.

### Supplemental Discussion

Our study sits at the intersection of medical imaging, advanced mathematical modeling, immunology, and oncology, focusing on the thymus; an organ whose role in adult health is only beginning to be fully appreciated. A recent publication by Kooshesh *et al.* in *The New England Journal of Medicine* linked adult thymectomy to increased risks of illness and mortality, stimulating broad interest in revisiting the long-standing view of the thymus as vestigial and inconsequential in adulthood.

For clinical oncologists, examining the implications of these findings for patients with cancer was a natural extension of this renewed interest, and multiple investigators within our center independently arrived at this topic. Thus, this work benefited from a deeply collaborative research environment. Because thymic structure and function are intrinsic and universal features of human biology, their description may necessarily converge across studies^3–5,7,8,10,18,21–24,34,38,39,41,46,47,83–87^. In the present study, we define a contouring atlas for the thymic region, introduce a novel context-aware automatic contouring framework for this structure, and present a robust, biologically grounded mathematical framework for quantifying persistent thymic tissue in patients with locally advanced NSCLC. The “percent thymic tissue” (pTT) metric builds on published, open-source concepts and methods, including, importantly, the quantitative framework first introduced by Okamura *et al.*, enabling standardized and reproducible assessment of thymic composition across diverse populations.

Our group is further developing a spatially aware extension of pTT and refining the framework to incorporate microvascular perfusion characteristics. Moreover, leveraging recent advances in interactive segmentation models^88^, we plan to integrate in-context learning strategies to further enhance the autocontouring framework’s performance (particularly in limited-data settings) for research purposes. Other groups at our center have independently developed clinically deployable thymus segmentation models; our complementary framework is intended for research use and emphasizes biological interpretability and translational insight. We are specifically validating pTT using a comprehensive repository of peripheral immune biomarkers, including circulating tumor markers, cytokines, and T-cell repertoire analyses, collected from patients with locally advanced NSCLC enrolled on institutional protocols. We anticipate that this work will establish pTT as the reference standard for non-invasive evaluation of thymic function in cancer, catalyzing research in this emerging domain and enabling future regulatory qualification and clinical adoption.

Moreover, in real-world clinical environments, particularly in resource-limited settings, artificial intelligence (AI) and deep-learning frameworks that depend on substantial GPU infrastructure or cloud-based compute resources may not be practical and are further limited by potential biases introduced through training on unbalanced datasets with underrepresentation of diverse populations. By contrast, pTT can be implemented on any standard workstation using readily available, inexpensive software. While the current proof-of-principle analysis automated thymic segmentation, the pTT metric itself is agnostic to segmentation technique and can be derived through manual or semi-automated contours with appropriate training. In an era when AI risks amplifying regional and racial disparities in cancer care^30^, pTT offers an interpretable, biologically grounded, scalable, and inherently unbiased, biomarker that is well positioned for equitable global deployment, pending further validation and review.

## Notes

### Competing Interest Statement

The authors have declared no competing interest.

### Funding Statement

This study was funded by NCI Cancer Center Support Grant (CCGS) P30 CA008748, and Radiological Society of North America (RSNA) #RR2311 (PI: Chaunzwa)

### Author Declarations

Ethics committee/IRB of Memorial Sloan Kettering Cancer Center, New York gave ethical approval for this work

